# Genomic surveillance at scale is required to detect newly emerging strains at an early timepoint

**DOI:** 10.1101/2021.01.12.21249613

**Authors:** Darcy Vavrek, Lucia Speroni, Kirsten J. Curnow, Michael Oberholzer, Vanessa Moeder, Phillip G. Febbo

## Abstract

Genomic surveillance in the setting of the coronavirus disease 2019 (COVID-19) pandemic has the potential to identify emerging SARS-CoV-2 strains that may be more transmissible, virulent, evade detection by standard diagnostic tests, or vaccine escapes. The rapid spread of the SARS-CoV-2 B.1.1.7 strain from southern England to other parts of the country and globe is a clear example of the impact of such strains. Early discovery of the B.1.1.7 strain was enabled through the proactive COVID-19 Genomics UK (COG-UK) program and the UK’s commitment to genomic surveillance, sequencing about 10% of positive samples.^1^ In order to enact more aggressive public health measures to minimize the spread of such strains, genomic surveillance needs to be of sufficient scale to detect early emergence and expansion in the broader virus population. By modeling common performance characteristics of available diagnostic and sequencing tests, we developed a model that assesses the sampling required to detect emerging strains when they are less than 1% of all strains in a population. This model demonstrates that 5% sampling of all positive tests allows the detection of emerging strains when they are a prevalence of 0.1% to 1.0%. While each country will determine their risk tolerance for the emergence of novel strains, as vaccines are distributed and we work to end the pandemic and prevent future SARS-CoV-2 outbreaks, genomic surveillance will be an integral part of success.

## Introduction

As it spreads across the globe and moves through communities, the SARS-CoV-2 virus is evolving at a rate of 1 to 2 variants per month.^2, 3^ Since the first full SARS-CoV-2 genome was published in January 2020^4^, sequencing of specimens from COVID-19 patients has allowed tracking of viral spread and identification of novel strains.^5-11^ Seminal work early in the pandemic demonstrated how SARS-CoV-2 had entered Washington state in the US^2^, with most early infections linked to a single introduction in late January or early February. In Northern California, analysis of 36 early samples demonstrated that there were multiple points of entry.^12^

With a full year of global transmission and over 89 million confirmed cases as of January 10, 2021,^13^ there have been 11 major clades identified.^14^ Importantly, specific strains that appear to be more transmissible have recently been identified in the UK^11, 15^ and South Africa.^10^ The emergence and growing predominance of these strains underscores the dynamic nature of the virus and highlights the importance of genomic surveillance. Of note, the US only began focused efforts to look for an emerging strain after the COG-UK program identified B.1.1.7, by which point the strain had already established itself within at least 6 states.^16^ Further spread of the more infections B.1.1.7 strain within the US, where many states already have overwhelmed health systems and ICU bed shortages, may have dramatic consequences.

As diagnostic capacity is scaled up and vaccines are distributed, systematic genomic surveillance will be a critical element to success in controlling the COVID-19 pandemic. Besides monitoring the SARS-CoV-2 virus population for changes in the biochemical properties underlying transmissibility, genomic epidemiology is critical to identify emergence of strains with potential to escape front line diagnostics, such as PCR and antigen tests.^17^ In addition, the vaccine can exert selective pressure and accelerate the emergence of potential lineages with altered antigenicity and potential reduced susceptibility to vaccine-induced immune response. Genomic surveillance has the potential to detect vaccine escape by emerging strains to enable a rapid response and potentially inform updates to current vaccine candidates. In this paper, a model is presented using assumptions from established diagnostic assays, to demonstrate the prevalence of a variant of concern (VOC) within positive diagnostic tests required to detect the early emergence of a novel strain. The specific amount of sampling will be determined by each community’s risk tolerance for missing strains that may be more transmissible, resistant to treatments, or vaccine escapes. Given the range of precedents with Australia working to sequence all positives samples^18^, the UK sequencing about 10%^1^, and other countries between 1% and 5%^19^, we chose 5% as an ambitious yet meaningful proportion. It is hoped that this model will provide a useful tool for public health entities.

## Methods

### Context

Presented in this paper is a model that describes detection of novel strains during SARS-CoV-2 screening as performed in the general population on a nationwide scale. As new viral mutations are identified in the population, some are determined to be different enough to be classified as a new variant, strain, and/or clade. Sometimes a new strain may be more transmissible and, in the setting of increasing distribution of vaccines, there may be vaccine escape strains (viral strains with mutation(s) that allows them to escape destruction by the host). In either case, it is relevant to identify new strains when they occur.

The scenario explored here is one where a percentage of the positive samples are sampled for sequencing to identify the viral strain. However, because SARS-COV-2 tests are not 100% accurate, false negatives can occur, some persons with the new strain may not be detected. It is also conceivable that all persons with the new strain will not be tested. For the purpose of this model, we focused on the prevalence of a new strain within test positive SARS-COV-2 samples. The prevalence within the surrounding population would need to be extrapolated.

There is also the potential for a false positive (FP) result, that the positive test sample is an error, and for this reason, only samples with a strong signal, e.g. cycle threshold (Ct) < 30, are recommended for further testing to identify the SARS-COV-2 strain in the sample. The models evaluated how detection of new variants was impacted by prevalence in the test positive sample population. The models can be used to estimate the minimum required prevalence of a new variant within the test positive samples selected for sequencing from the surrounding population.

### Testing Overview and Model Assumptions

The model is based upon reflex sequencing of diagnostic samples found to be positive with the following assumptions: Persons are tested for SARS-CoV-2 nationwide with minimal introduction of bias; approximately 80% of true positive (TP) cases, and no FP cases, will have a Ct < 30;^20^ prevalence of the surrounding population will impact the number of FP vs TP results; it is of interest to determine the required prevalence of a new strain within TP samples in order to be detected.

### Metrics to Evaluate

The following metrics and dependencies were set or determined in the development and running of the model:

1. Expected number of positive samples (Npos) out of N tested, which depends upon prevalence within the samples collected and screened, test sensitivity, and test specificity.
2. Expected number of true positive (TP) SARS-CoV-2 samples within NPos, which also depends upon the sensitivity and specificity of the screening test utilized.
  - In the US population (nationwide) and 1/50^th^ of the US population (state level with equal split by state), 14% prevalence, 80% screening sensitivity, and 99% screening specificity were assumed.
  - In Los Angeles (LA) County, California and Dane County, Wisconsin test we used recently reported sample test volume numbers and test positivity rates. These locations are likely using several different tests, which may be of varying performance. To address this, we used two different performance combinations to generate a broad range of screening sample prevalences that would explain observed test positivity results: 1, high specificity performance, with an 80% screening sensitivity and 99% screening specificity; 2, high sensitivity performance, with 90% screening sensitivity and 95% screening specificity. This allows for the calculation of higher and lower estimates of reflex sequencing sample volume to enter into modeling.
3. Expected number of TPs with a strong enough signal for reflex sequencing. It is assumed, for modeling purposes, that 80% of the TP samples will have a strong enough viral load (Ct < 30) to be candidates for reflex sequencing. Conversely, it is assumed that none of the false positive samples would have a strong enough viral load to be candidates for reflexing.
4. Expected selection rate of qualifying samples is 5%. If approximately 5% of TP samples with a strong viral load will be sequenced, RNT (Reflex sequencing Number Tested) sequenced samples will also have an additional contingency table with respect to those samples that have the VOC versus those that do not. Sensitivity and specificity to detect the new strain with reflex sequencing when present is assumed to be 80% and 99%.
5. Expected number of positive results within samples selected for RNT depends upon the prevalence of the VOC within the selected samples (modeling assumes a range of 0.005% to 10%).

### Mathematical Modeling and Formulae

The models presented used the formulae presented in Tables 1-4. Terms used in the model formulae and contingency tables are presented in Table 1. These terms can also be summarized in contingency tables which are presented to describe test results versus comparator results (Table 2). This will be done first in screening samples with respect to the SARS-CoV-2 infected status of the person tested (Table 3). The formulae used to fill out Table 3 demonstrates that all numbers of the table can be derived from the total number of samples screened, prevalence within screened samples, and the sensitivity and specificity of the assay. The next set of formulae, Table 4, shows that all numbers of the table can be derived from the total number of samples selected for reflex sequencing (RNT), the prevalence of the novel strain within tested samples, and the sensitivity and specificity of the assay used for reflex sequencing, similarly to Table 3.

**Table 1:** Definitions of terms used in Formulae.

| Term | Definition |
| --- | --- |
| N | The number of persons screened in the population |
| prev;<br>prev.rt | Prevalence (prev) is the percentage of infected samples among those who are screened. Prev.rt is the percentage of reflex sequencing tested samples infected with a novel strain. |
| sens;<br>sens.rt | Sensitivity (sens) is the probability of a screen test positive result in a truly infected sample. Sens.rt is the sensitivity of the test used for sequencing the virus. |
| spec;<br>spec.rt | Specificity is the probability of a screen test negative result in a sample which is truly not infected. Spec.rt is the specificity of the test used for sequencing the virus. |
| TP;<br>TP.screen | True Positive (TP) is a test positive result in an infected sample. TP.screen are the screened positive results that are then considered for reflex sequencing. |
| FP | False Positive (FP) is a test positive result in a sample which is not infected |
| TN | True Negative (TN) is a test negative result in a sample which is not infected |
| FN | False Negative (FN) is a test negative result in an infected sample |
| Npos | The number test positive (Npos) is the number of test positive samples |
| Nneg | The number test negative (Nneg) is the number of test negative samples |
| Ninf | Number infected (Ninf) is the number of infected samples |
| NH | Number healthy (NH) is the number of samples which are not infected |
| ctstrong | The percentage of screen positive, true positive results with a strong enough viral load (ct < 30) to be sequenced |
| selected | The percentage of screen test positive samples with a strong viral load which are selected for reflex sequencing |
| RNT | Reflex sequencing Number Tested (RNT) is the number of eligible samples selected for sequencing |

**Table 2:** Contingency table of test results versus comparator results.

|  | **Comparator Outcome** |  |  |
| --- | --- | --- | --- |
| **Assay Result** | **Present** | **Not present** | **Total** |
| **Test positive** | True Positive (TP) | False Positive (FP) | Npos |
| **Test negative** | False Negative (FN) | True Negative (TN) | Nneg |
| **Total** | Number infected (Ninf) | Number healthy or not infected (NH) | Number of samples (N) |

**Table 3:**
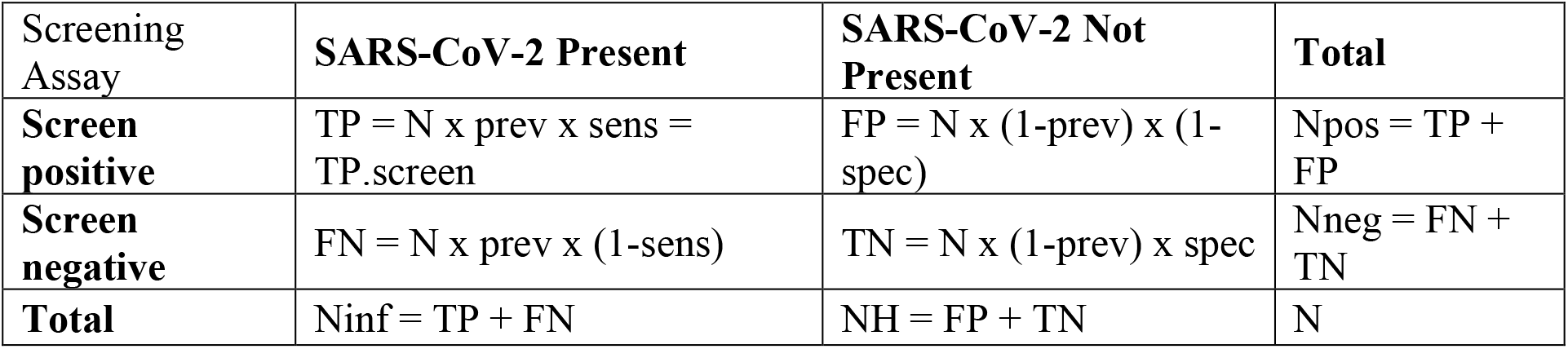
Contingency table formulae for screening test results versus SARS-CoV-2 status.

**Table 4:** Contingency table formulae of reflex sequencing test results versus comparator results.

| Reflex Sequencing | **SARS-CoV-2 VOC PRESENT** | **SARS-CoV-2 VOC NOT PRESENT** | **Total** |
| --- | --- | --- | --- |
| **Positive for VOC** | $TP = RNT \times \text{prev.rt} \times \text{sens.rt}$ | $FP = RNT \times (1 - \text{prev.rt}) \times (1 - \text{spec.rt})$ | $N_{\text{pos}} = TP + FP$ |
| **Negative for VOC** | $FN = RNT \times \text{prev.rt} \times (1 - \text{sens.rt})$ | $TN = RNT \times (1 - \text{prev.rt}) \times \text{spec.rt}$ | $N_{\text{neg}} = FN + TN$ |
| **Total** | $N_{\text{inf}} = TP + FN$ | $NH = FP + TN$ | $RNT = TP_{\text{screen}} \times \text{ctstrong} \times \text{selected}$ |

VOC, variant of concern.

Calculations related to reflex sequencing were made across a range of prevalences in order to demonstrate the prevalence necessary within the reflex sequencing population of samples in order to be detected. To avoid presenting numerous summary contingency tables, these results will be shown graphically in this paper, with expected true positive results on the y-axis with respect to prevalence of the new strain within reflex sequenced samples on the x-axis.

To assess expected sample volume to enter into the models, estimates were made with respect to daily screening tests within the US population to get a sense of the potential volume of true positive screen samples that would be eligible for reflex sequencing, and the prevalence necessary for detection of the VOC. This is followed by numbers that are 1/50^th^ the size of national daily screening (as an estimate of state level numbers), by weekly screening tests within LA County, and finally with weekly screening numbers in Dane County.

The LA County and Dane County locations were selected to show the implications of the model using current testing numbers. The LA County location was chosen as an example of a large population experiencing a current outbreak. The Dane County, Wisconsin location was chosen to reflect a smaller population with a high current test positive rate. When estimating sample volumes in LA County, California and Dane County, Wisconsin different screening sample prevalences and screening test performances were assumed to get a range for the sample volume that might be expected weekly for reflex sequencing and that are consistent with weekly reported number of tests and test positivity rates in these locations.

## Results

### Positive Diagnostic Test Samples Available to Sequence

The number of positive diagnostic tests for SARS-COV-2 within the population represents the sample pool from which sequencing is performed to detect a novel strain, or VOC. National daily testing in the US has been between 1 million (M) and 1.5M since early October^21^ and percent positive results have varied but herein is modeled as 14%. Here, we have used a value of 1M samples tested daily, and with the assumptions of diagnostic test performance (80% sensitivity, 99% specificity; a minimum sensitivity of 80% is consistent with antigen assays^22^), this results in 120,600 positive tests, of which 112,000 are expected to be true positive (Table 5). If 80% of these true positive samples have a high enough viral load signal for retesting (e.g. Ct < 30), 89,600 SARS-COV-2 samples (112,000 x 80% = 89,600) are candidates for daily surveillance for a new strain.

**Table 5:** **Cross-tabulation of expected SARS-CoV-2 test results versus the true status in 1 million persons tested daily based on 14% prevalence within tested US population, 80% test sensitivity, and 99% test specificity**

| Screening Assay | SARS-CoV-2 Present | SARS-CoV-2 Not Present | Total |
| --- | --- | --- | --- |
| **Screen Positive** | 112,000 | 8,600 | 120,600 |
| **Screen Negative** | 28,000 | 851,400 | 879,400 |
| **Total** | 140,000 | 860,000 | 1,000,000 |

### Sequencing of Positive Samples

If we take those 89,600 candidate TP samples, and select 5% for reflex sequencing, that would be 4,480 samples nationwide for daily surveillance. Table 6 provides the expected results for this nationwide testing based on a 1% prevalence of the VOC. Table 7 provides the expected result based at a state level if samples were assumed to be equally distributed per state (not a population-based distribution), i.e., 4,480 samples / 50 states = 90 samples per state. Sampling over time in a population may result in different potential sample volumes for detecting a VOC as depicted in Figure 1. Importantly, regardless of sampling rate from eligible samples, detection of VOC does not appreciably occur prior to the VOC achieving 0.1% prevalence. The models presented assume that the samples selected for reflex sequencing are chosen without bias and are representative of both the greater population of eligible samples for testing and of patients screened for SARS-CoV-2. With proper sampling, the volume for reflex sequencing in a state with less positive screen samples available still provides enough test samples for VOC when the prevalence rises above 0.1% (Figure 2). Therefore, while changes in modeling assumptions clearly impact the prevalence at which a VOC will be detected with some confidence, our model assumption sampling rate of 5% of eligible positives is likely sufficient to detect an emerging VOC at a prevalence between 0.1% and 1.0%.

**Table 6:** **Detection of variants of concern through daily reflex sequencing nationwide. Cross tabulation of expected daily reflex sequencing test result versus true status in 4**,**480 selected screen positive samples with a strong viral load tested based on 1% prevalence within reflex sequenced US population, 80% reflex sequencing sensitivity, and 99% reflex sequencing specificity**.

| Reflex Sequencing | SARS-CoV-2 VOC<br>PRESENT | SARS-CoV-2 VOC<br>NOT PRESENT | Total |
| --- | --- | --- | --- |
| **Positive for VOC** | 36 | 44 | 80 |
| **Negative for VOC** | 9 | 4,391 | 4,400 |
| **Total** | 45 | 4,435 | 4,480 |

VOC, variant of concern.

**Table 7:** **Detection of variants of concern at a state level. Cross tabulation of expected daily, in a given state, reflex sequencing test result versus true status in 90 positive samples with a strong viral load tested based on 1% prevalence within reflex sequenced 1/50**^**th**^ **of the US population, 80% reflex sequencing sensitivity, and 99% reflex sequencing specificity**

| Reflex Sequencing | SARS-CoV-2 VOC<br>PRESENT | SARS-CoV-2 VOC<br>NOT PRESENT | Total |
| --- | --- | --- | --- |
| **Positive for VOC** | 1 | 1 | 2 |
| **Negative for VOC** | 0 | 88 | 88 |
| **Total** | 1 | 89 | 90 |

VOC, variant of concern.

**Figure 1.**
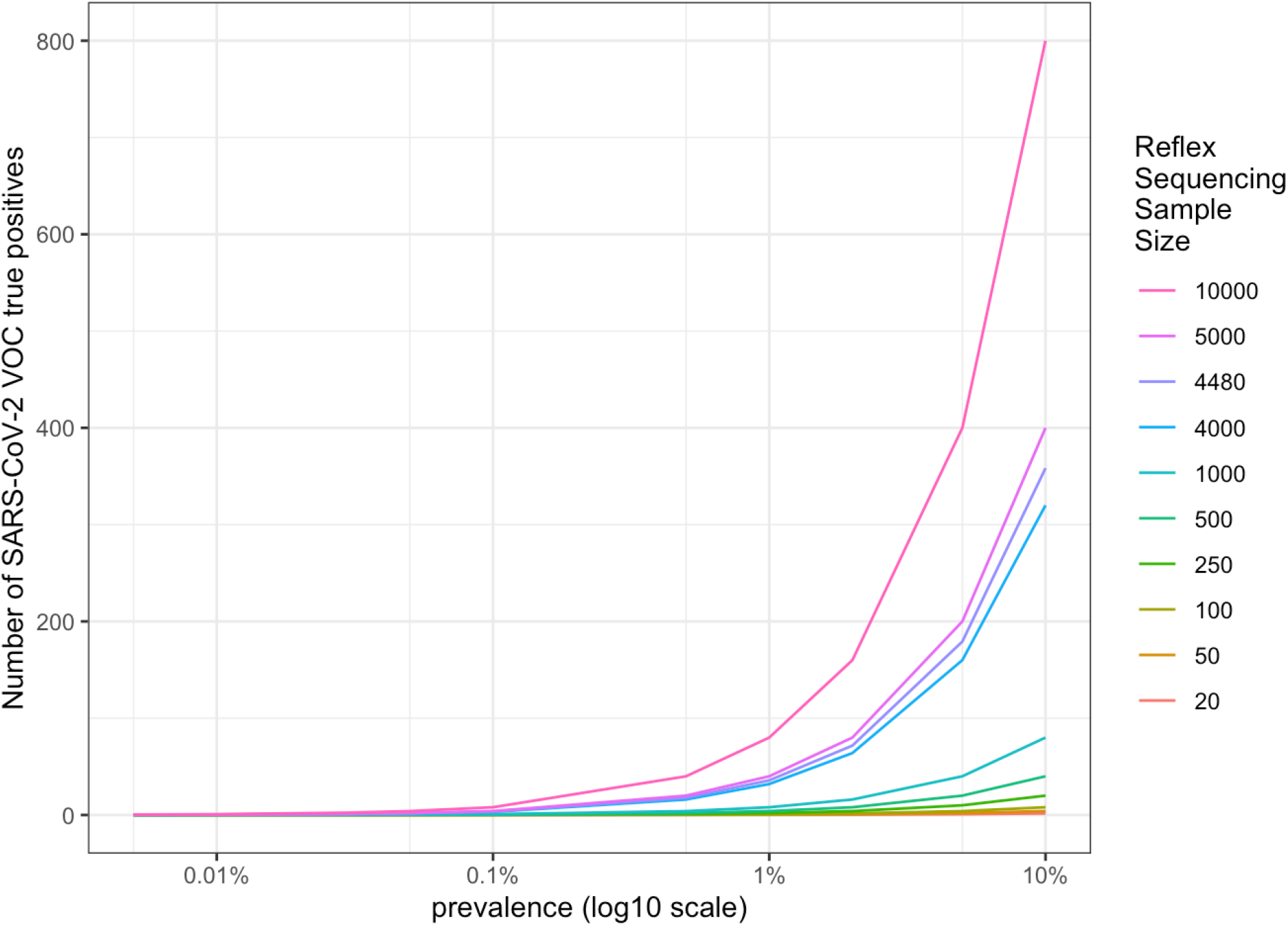
**Number of true positive samples when detecting SARS-COV-2 VOC amongst a sample of reflex sequenced screened positive samples with a strong viral load, based upon prevalence of the VOC within larger reflex sequenced sample of US population**

**Figure 2.**
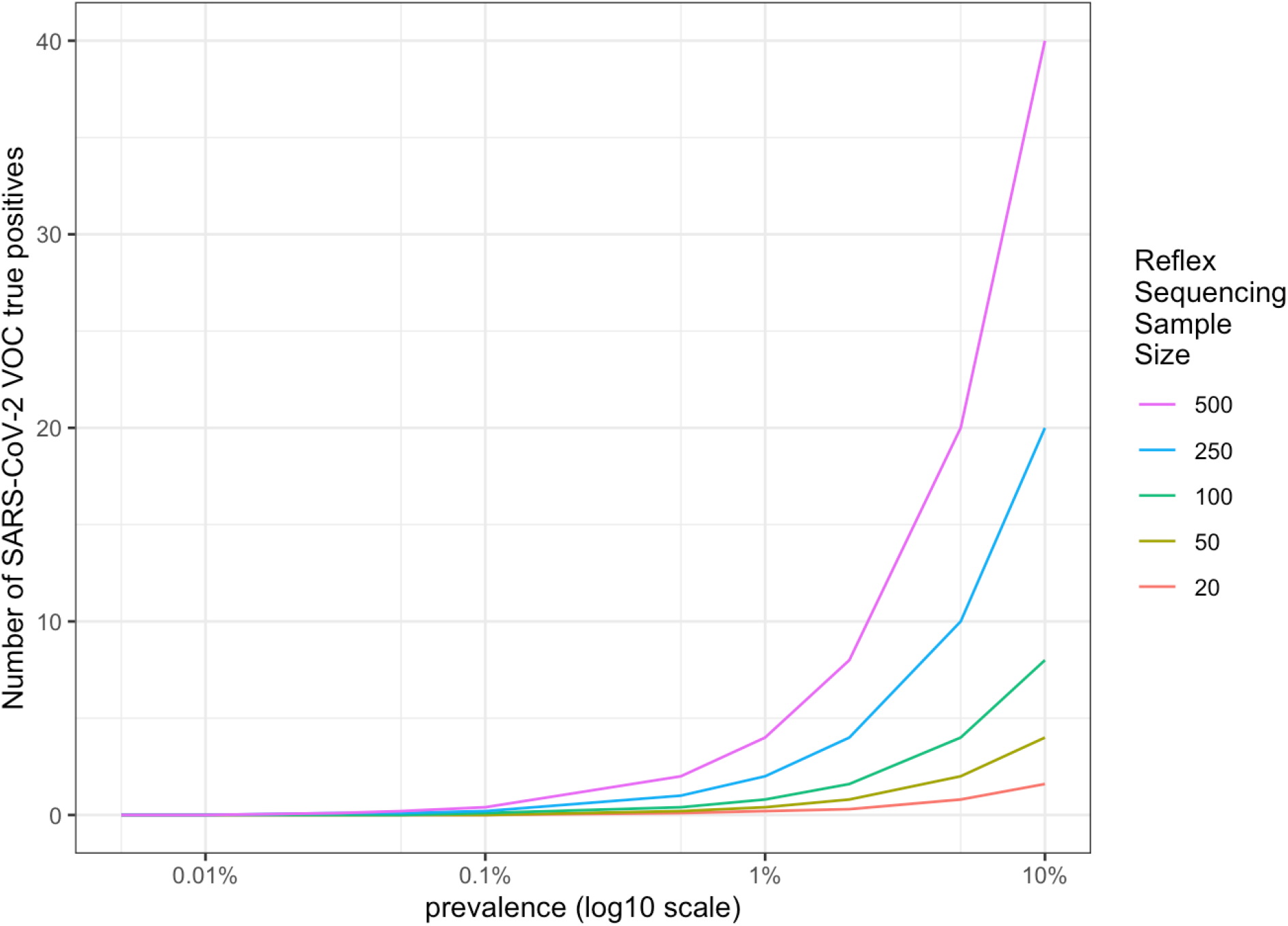
Number of true positive samples when detecting SARS-COV-2 VOC. amongst a sample of reflex sequenced screened positive samples with a strong viral load, based upon prevalence of the VOC within smaller reflex sequenced sample of 1/50th of US populations

As state and county sampling are more likely to be population based, we looked at the implications of the model in two US counties using specific numbers that are available in those locations.

### Model applied to LA County

We applied the model to LA County where approximately 300,000 tests are performed per day with a positive testing rate of 10%, which is around 30,000 positive tests per day. There are different diagnostic tests being utilized, but if the aggregate test performance is 80% sensitivity and 99% specificity, these LA County numbers would indicate a prevalence of 11.5% (Table 8). However, if the average screening sensitivity was higher at 90%, but the average screening specificity was lower at 95%, this would indicate a prevalence of 6% (Table 9). Based on the 300,000 samples per day, we assumed 2 million samples tested per week, with approximately 200,000 screen positives. Of the 200,000 screen positive samples, 100,000 to 180,000 would be true positives and 80,000 to 144,000 of these would have a high enough viral load for reflex sequencing (80% of the true positives). Sequencing 5% of these 80,000-114,000 screen positive samples with a high viral load, gives 4,000 to 7,200 samples per week undergoing reflex sequencing and would result in a VOC being detected when the prevalence within retested samples is greater than 0.1% (Figure 3).

**Table 8:**
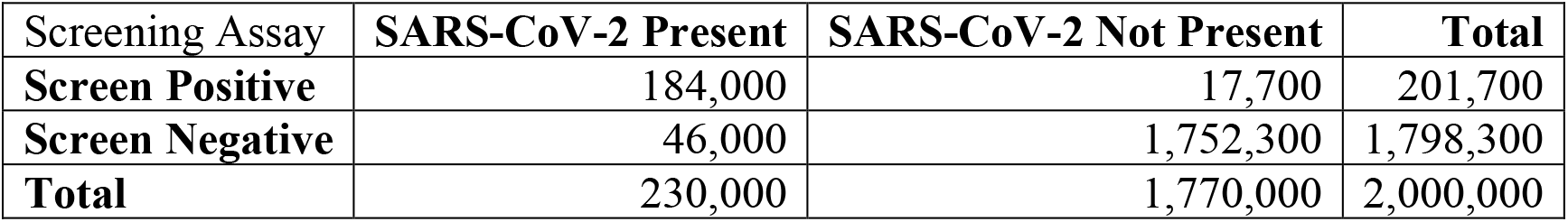
**Cross-tabulation of expected SARS-CoV-2 screening test results versus the true status in 2 million persons tested weekly in LA County, California based on 11**.**5% prevalence within tested population, 80% test sensitivity, and 99% test specificity**

| Screening Assay | SARS-CoV-2 Present | SARS-CoV-2 Not Present | Total |
| --- | --- | --- | --- |
| **Screen Positive** | 184,000 | 17,700 | 201,700 |
| **Screen Negative** | 46,000 | 1,752,300 | 1,798,300 |
| **Total** | 230,000 | 1,770,000 | 2,000,000 |

**Table 9:** **Cross-tabulation of expected SARS-CoV-2 screening test results versus the true status in 2 million persons tested weekly in LA County, California based on 6% prevalence within tested population, 90% test sensitivity, and 95% test specificity**

| Screening Assay | SARS-CoV-2 Present | SARS-CoV-2 Not Present | Total |
| --- | --- | --- | --- |
| **Screen Positive** | 108,000 | 94,000 | 202,000 |
| **Screen Negative** | 12,000 | 1,786,000 | 1,798,000 |
| **Total** | 120,000 | 1,880,000 | 2,000,000 |

**Figure 3:**
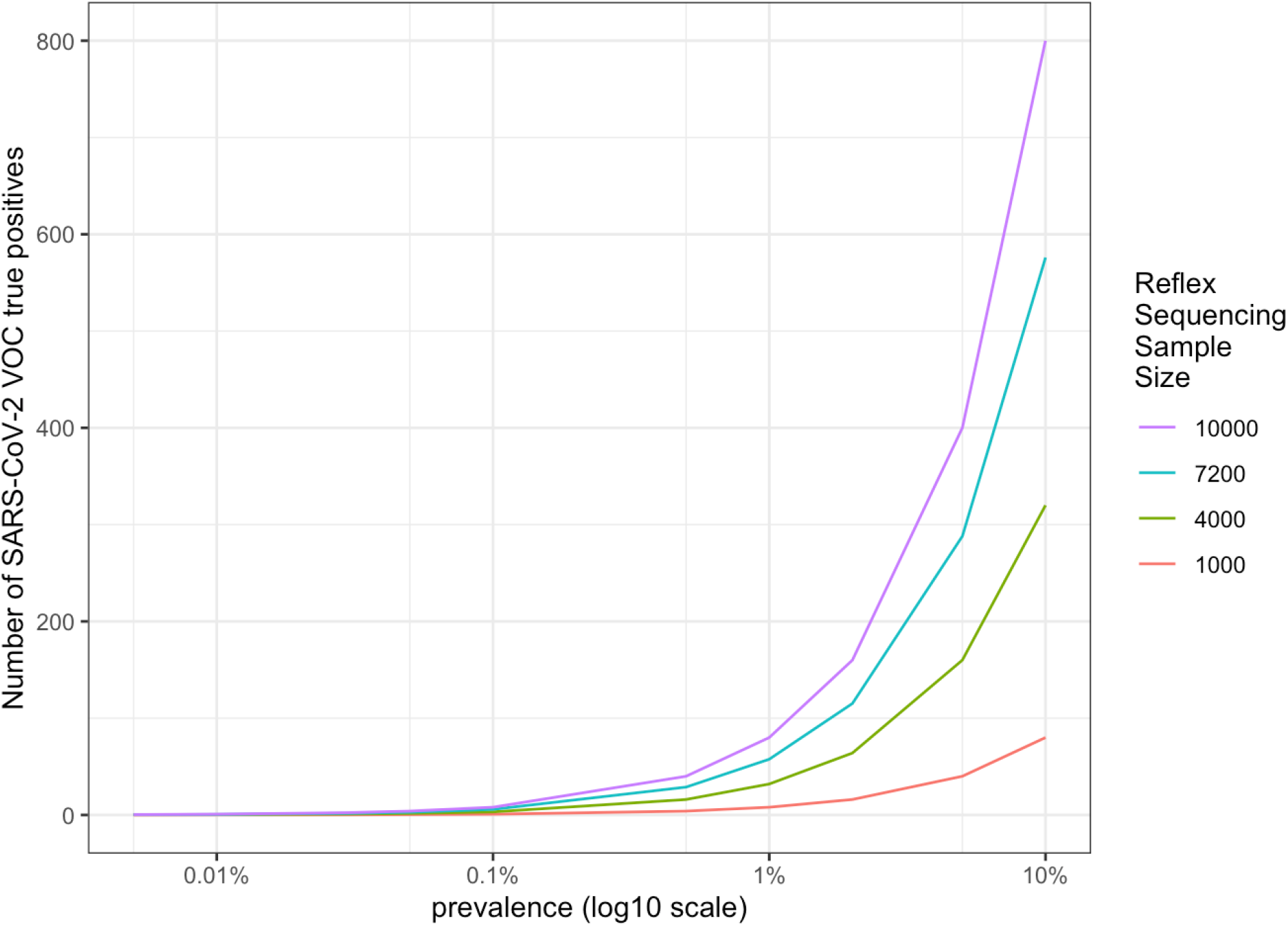
**Number of true positive samples when detecting SARS-COV-2 VOC amongst a sample of reflex sequenced screened positive samples with a strong viral load, based upon prevalence of the VOC within smaller reflex sequenced sample of LA County, California population**

### Model applied to Dane County, Wisconsin

For Dane county, we assumed approximately 7,000 to 12,000 tests per day or 75,000 tests per week. An estimate of 2,000-4,000 positive tests per day or 25,000 per week (high estimate) could be explained by a screening population prevalence of 41% with test performance of 80% sensitivity and 99% specificity (Table 10). However, the number of screened TPs may be slightly less if the prevalence is lower at 34%, specificity is lower 95%, and sensitivity is higher 90% amongst tested cases (Table 11). The range of expected samples in Dane County that are true positive is estimated to be 22,000 to 25,000 of the 75,000 weekly samples, 17,600 to 20,000 of these true positives would have high enough viral load for retesting, and 5% of this is 880 to 1000 samples per week for reflex sequencing. True positive samples start getting detected when the prevalence within retested samples is greater than 0.1% (Figure 4).

**Table 10:** **Cross-tabulation of expected SARS-CoV-2 screening test results versus the true status in 75**,**000 persons tested weekly in Dane County, Wisconsin based on 41% test prevalence within tested population, 80% test sensitivity, and 99% test specificity**

| Screening Assay | SARS-CoV-2 Present | SARS-CoV-2 Not Present | Total |
| --- | --- | --- | --- |
| **Screen Positive** | 24,600 | 443 | 25,042 |
| **Screen Negative** | 6,150 | 43,808 | 49,958 |
| **Total** | 30,750 | 44,250 | 75,000 |

**Table 11:** **Cross-tabulation of expected SARS-CoV-2 screening test results versus the true status in 75**,**000 persons tested weekly in Dane County, Wisconsin based on 34% test prevalence within tested population, 90% test sensitivity, and 95% test specificity**

| Screening Assay | SARS-CoV-2 Present | SARS-CoV-2 Not Present | Total |
| --- | --- | --- | --- |
| **Screen Positive** | 22,950 | 2,475 | 25,425 |
| **Screen Negative** | 2,550 | 47,025 | 49,575 |
| **Total** | 25,500 | 49,500 | 75,000 |

**Figure 4:**
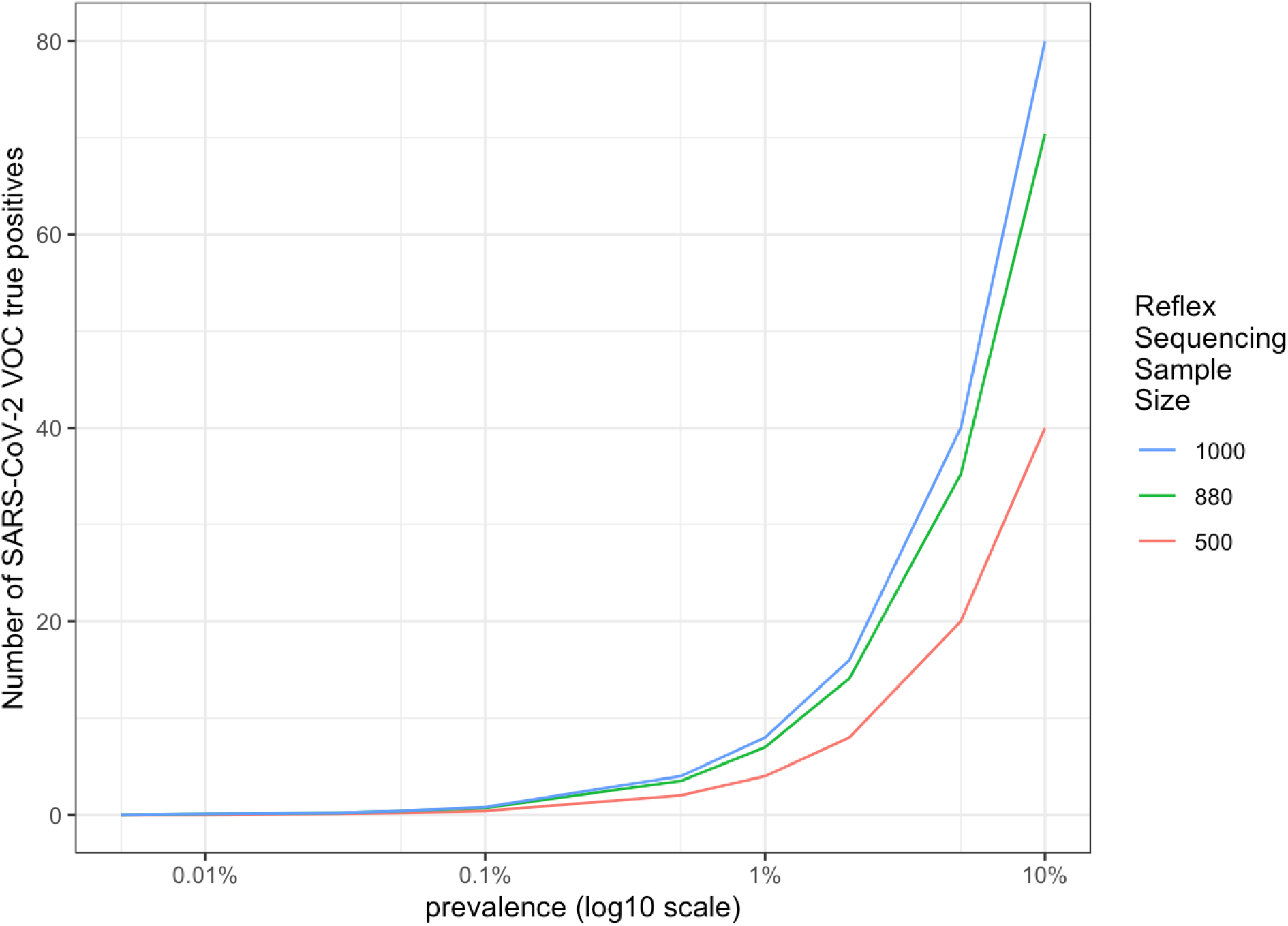
**Number of true positive samples when detecting SARS-COV-2 VOC amongst a sample of reflex sequenced screened positive samples with a strong viral load, based upon prevalence of the VOC within smaller reflex sequenced sample of Dane County, Wisconsin population**

## Discussion

We begin 2021 with the paradox of an escalating number of cases, hospitalized patients, and record level mortality due to COVID-19 while at the same time there is the distribution of highly effective vaccines, robust diagnostic testing, and improved management of those who do become sick. Genomic surveillance is underutilized in the United States despite the broad availability of sequencing technology and the existence of capable public, private, and academic laboratories of ample scale that can be utilized to develop a robust systematic, nationwide genomic surveillance program.

The utilization of genomic surveillance globally varies quite significantly between different countries. Some countries, like Australia, aim to sequence all SARS-COV-2 positive samples and use this genomic surveillance information to rapidly identify SARS-COV-2 transmission chains and to guide their public health responses^6^, such as localized stay-at-home orders and travel restrictions (i.e., limiting incoming travel numbers and triggering within country border closures). For Australia, the smaller population size and relatively low number of SARS-COV-2 cases make a comprehensive genomic surveillance approach easier to implement. Other countries, like the US and less developed nations, have limited current utilization of genomic surveillance. In the US, the large population and current high prevalence of SARS-COV-2 makes genomic profiling of all SARS-COV-2 positive samples impractical from a logistical and cost perspective. Our modeling suggests that getting to a 5% sampling of positive cases for reflex sequencing will allow detection of emerging VOCs with a prevalence of 0.1% to 1.0% within a community. This allows public health officials to move quickly to institute appropriate measures to better contain such variants. As we have seen in the UK, once the variant achieves greater than 1% prevalence, there is little opportunity for containment. Further, rapid identification of emerging VOCs would allow researchers to quickly act to modify diagnostics, therapeutics, and vaccines.

Vaccines for COVID-19 have been developed and approved in record time. While there is a push to get the global population vaccinated as quickly as possible, it will take at least 6 months, and likely closer to a year, before the broader population is vaccinated. The primary limitation is vaccine availability, and although vaccine developers are ramping up supply as quickly as possible, it will take time to produce the huge volume of doses needed. Further, safety and efficacy in some populations, such as children, has not yet been established. As vaccines are delivered, there is now a selective pressure and risk for vaccine escape variants. Immune escape variants have been recently reported in patients treated for COVID-19 infection.^23, 24^ Thus, genomic surveillance for vaccine escapes is now critically important as undetected emergence of a vaccine escape strain could hinder the ability to end this pandemic. As vaccination timelines will vary between countries, it will remain important to continue surveillance for these variants even once a country is fully vaccinated as there could be introductions from unvaccinated individuals travelling from countries with lower vaccination rates.

Our model is based on many assumptions that may or may not be valid for specific regions and/or countries. We also make assumptions that are idealistic such as minimized sampling bias with similar distributions of any relevant sample, patient, and viral characteristics across populations. As with any model, it is imperfect. It is proposed to help demonstrate the value that sequencing just 5% of positive samples provides to public health officials and other stakeholders in order to set a goal for genomic surveillance. Genomic surveillance, based on sequencing of positive COVID-19 samples, can only report on strains detected by standard of care PCRs. As strains evolve, inclusion of samples with undetermined PCR test result may be advantageous to further monitor for strains which escape diagnostic tests on the market.

While it is anticipated that the number of confirmed cases of SARS-COV-2 will decrease as an increasing proportion of the population is vaccinated, genomic surveillance becomes even more important in post-vaccinated individuals and populations due to the likely emergence of vaccine escape strains. Vaccines can be modified and adapted when resistant strains are identified, but vaccine modifications take time to develop and test. Thus, the earlier such strains can be identified and contained, the more likely we are to truly end the pandemic.

## Data Availability

Questions on data availability can be directed to the corresponding author, Darcy Vavrek

## References

1. COVID-19 Genomics UK Consortium (COG-UK). Report on SARS-CoV-2 Spike mutations of special interest. December 20, 2020. Accessed January 11, 2021. https://www.cogconsortium.uk/wp-content/uploads/2020/12/Report-1_COG-UK_20-Decmber-2020_SARS-CoV-2-Mutations_final_updated2.pdf

2. Bedford T, Greninger AL, Roychoudhury P, et al. Cryptic transmission of SARS-CoV-2 in Washington state. Science. Oct 30 2020;370(6516):571–575. doi:10.1126/science.abc0523

3. Rambaut A. Phylogenetic analysis of nCoV-2019 genomes. Accessed January 10, 2021. https://virological.org/t/phylodynamic-analysis-176-genomes-6-mar-2020/356

4. Zhu N, Zhang D, Wang W, et al. A Novel Coronavirus from Patients with Pneumonia in China, 2019. N Engl J Med. Feb 20 2020;382(8):727–733. doi:10.1056/NEJMoa2001017

5. Gudbjartsson DF, Helgason A, Jonsson H, et al. Spread of SARS-CoV-2 in the Icelandic Population. N Engl J Med. Jun 11 2020;382(24):2302–2315. doi:10.1056/NEJMoa2006100

6. Seemann T, Lane CR, Sherry NL, et al. Tracking the COVID-19 pandemic in Australia using genomics. Nat Commun. Sep 1 2020;11(1):4376. doi:10.1038/s41467-020-18314-x

7. Oude Munnink BB, Nieuwenhuijse DF, Stein M, et al. Rapid SARS-CoV-2 whole-genome sequencing and analysis for informed public health decision-making in the Netherlands. Nat Med. Sep 2020;26(9):1405–1410. doi:10.1038/s41591-020-0997-y

8. Page AJ, Mather AE, Le-Viet T, et al. Large scale sequencing of SARS-CoV-2 genomes from one region allows detailed epidemiology and enables local outbreak management. medRxiv. 2020:2020.09.28.20201475. doi:10.1101/2020.09.28.20201475

9. Lemieux JE, Siddle KJ, Shaw BM, et al. Phylogenetic analysis of SARS-CoV-2 in Boston highlights the impact of superspreading events. Science. Dec 10 2020;doi:10.1126/science.abe3261

10. Tegally H, Wilkinson E, Giovanetti M, et al. Emergence and rapid spread of a new severe acute respiratory syndrome-related coronavirus 2 (SARS-CoV-2) lineage with multiple spike mutations in South Africa. medRxiv. 2020:2020.12.21.20248640. doi:10.1101/2020.12.21.20248640

11. Volz E, Mishra S, Chand M, et al. Transmission of SARS-CoV-2 Lineage B.1.1.7 in England: Insights from linking epidemiological and genetic data. medRxiv. 2021:2020.12.30.20249034. doi:10.1101/2020.12.30.20249034

12. Deng X, Gu W, Federman S, et al. Genomic surveillance reveals multiple introductions of SARS-CoV-2 into Northern California. Science. Jul 31 2020;369(6503):582–587. doi:10.1126/science.abb9263

13. Johns Hopkins University, Center for Systems Science and Engineering (CSSE). COVID-19 Dashboard. Accessed January 10, 2021. https://coronavirus.jhu.edu/map.html

14. Nextstrain. Accessed January 10, 2021. https://nextstrain.org/staging/ncov/global/clades-update

15. Rambaut AL, N., Pybus, O., Barclay, W., Barrett, J., Carabelli, A., Connor, T., Peacock, T., Robertson, D.L., Volz, E., on behalf of COVID-19 Genomics Consortium UK (CoG-UK). Preliminary genomic characterisation of an emergent SARS-CoV-2 lineage in the UK defined by a novel set of spike mutations. Accessed January 10, 2021. https://virological.org/t/563

16. Illumina and Helix Collaborate to Assess Prevalence of New SARS-CoV-2 UK Variant (B.1.1.7) in the US and Develop National Surveillance Infrastructure. January 5, 2021, Accessed January 10, 2021. https://www.illumina.com/company/news-center/press-releases/press-release-details.html?newsid=ba453369-4367-424f-8f5d-eacdff3ca5f0

17. United States Food and Drug Administration. Genetic Variants of SARS-CoV-2 May Lead to False Negative Results with Molecular Tests for Detection of SARS-CoV-2 -Letter to Clinical Laboratory Staff and Health Care Providers. January 8, 2021, Accessed January 11, 2021. https://www.fda.gov/medical-devices/letters-health-care-providers/genetic-variants-sars-cov-2-may-lead-false-negative-results-molecular-tests-detection-sars-cov-2

18. Australian Government, Department of Health. Australian Health Protection Principal Committee (AHPPC) Statement on management of COVID-19 variants. January 8, 2021, Accessed January 12, 2021. https://www.health.gov.au/news/australian-health-protection-principal-committee-ahppc-statement-on-management-of-covid-19-variants

19. Stevens H. U.S. ranks 43rd worldwide in sequencing to check for coronavirus variants like the one found in the U.K. The Washington Post. December 23, 2020. Accessed January 12, 2021. https://www.washingtonpost.com/world/2020/12/23/us-leads-world-coronavirus-cases-ranks-43rd-sequencing-check-variants/

20. Lu J. (President, Helix, San Mateo, CA). Communication of Helix Diagnostics testing results.

21. Johns Hopkins University. Coronavirus Resource Center (CRC). Daily State-By-State Testing Trends. Accessed January 10, 2021. https://coronavirus.jhu.edu/testing/individual-states

22. FIND. Evaluation of SARS-CoV-2 Antigen (Ag) Detecting Tests. Accessed January 12, 2021. https://www.finddx.org/covid-19-old/sarscov2-eval-antigen/

23. Choi B, Choudhary MC, Regan J, et al. Persistence and Evolution of SARS-CoV-2 in an Immunocompromised Host. N Engl J Med. Dec 3 2020;383(23):2291–2293. doi:10.1056/NEJMc2031364

24. Kemp SA, Collier DA, Datir R, et al. Neutralising antibodies in Spike mediated SARS-CoV-2 adaptation. MedRxiv. 2020:2020.12.05.20241927. doi:10.1101/2020.12.05.20241927

